# Clinical conditions and their impact on utility of genetic scores for prediction of acute coronary syndrome

**DOI:** 10.1101/2020.09.16.20195883

**Authors:** Jiwoo Lee, Tuomo Kiiskinen, Nina Mars, Sakari Jukarainen, FinnGen Project, Erik Ingelsson, Benjamin Neale, Samuli Ripatti, Pradeep Natarajan, Andrea Ganna

## Abstract

**Aims:** Early prediction of acute coronary syndrome (ACS) is a major goal for prevention of coronary heart disease (CHD). Genetic information has been proposed to improve prediction beyond well-established clinical risk factors. While polygenic scores (PS) can capture an individual’s genetic risk for ACS, its prediction performance may vary in the context of diverse correlated clinical conditions. Here, we aimed to test whether clinical conditions impact the association between PS and ACS.

**Methods and Results:** We explored the association between 405 clinical conditions diagnosed before baseline and 9,080 incident cases of ACS in 387,832 individuals from the UK Biobank. We identified 80 conventional (e.g., stable angina pectoris (SAP), type 2 diabetes mellitus) and unconventional (e.g., diaphragmatic hernia, inguinal hernia) associations with ACS. Results were replicated in 6,430 incident cases of ACS in 177,876 individuals from FinnGen. The association between PS and ACS was consistent in individuals with and without most clinical conditions. However, a diagnosis of SAP yielded a differential association between PS and ACS. PS was associated with a significantly reduced (interaction p-value=2.87×10^−8^) risk for ACS in individuals with SAP (HR=1.163 [95% CI: 1.082–1.251]) compared to individuals without SAP (HR=1.531 [95% CI: 1.497–1.565]). These findings were replicated in FinnGen (interaction p-value=1.38×10^−6^).

**Conclusion:** In summary, while most clinical conditions did not impact utility of PS for prediction of ACS, we found that PS was substantially less predictive of ACS in individuals with prevalent stable CHD. PS for ACS may be more appropriate for asymptomatic individuals than symptomatic individuals with clinical suspicion for CHD.

## Introduction

Acute coronary syndrome (ACS), including myocardial infarction, is an unstable consequence of coronary heart disease (CHD), the leading cause of death worldwide. In Europe, CHD accounts for 1.8 million deaths per year, making up almost 20% of all cardiovascular disease-related deaths (1). Thus, prediction of ACS remains a major public health issue for prevention of CHD.

Well-established clinical risk factors, such as type 2 diabetes mellitus and hypercholesterolemia, are highly predictive of future health outcomes, including ACS (5, 6). Individuals with an accumulation of such risk factors are candidates for preventative statin interventions per American and European guidelines (5-7). Recently, other risk-enhancing factors independently associated with ACS have been proposed to improve prediction of ACS and inform statin intervention decisions (8). An unbiased, comprehensive evaluation of diverse correlated clinical conditions in large-scale biobanks may better inform prediction of ACS (9-11).

Furthermore, genetic information may inform prediction of ACS beyond well-established clinical risk factors (12-16). Currently, guidelines on evaluating risk for ACS and initiating statin interventions rely on several clinical risk factors but do not support the use of genetic information. However, genetic information has the advantage of remaining stable throughout life and therefore could be used to improve earlier prediction of ACS (17-19). Results from large genome-wide association studies (GWAS) can be used to derive polygenic scores (PS) based on millions of single nucleotide polymorphisms (SNPs) that are robustly associated with CHD (20). Several studies have investigated the use of PS for prediction of ACS in addition to well-established clinical risk factors (16, 21-23). However, there are no studies to date that examine the association between PS and ACS in the context of diverse correlated clinical conditions.

In this study, we have two main goals that we assessed in two large-scale biobanks. First, we aimed to comprehensively explore the association of clinical conditions with ACS. Second, we aimed to assess whether clinical conditions associated with ACS impact the association between PS and ACS to provide context for the use of PS in a clinical setting for prediction of ACS.

## Methods

### Study population

This study utilized data from the UK Biobank and replicated findings in FinnGen. The UK Biobank is a prospective cohort study that includes approximately 500,000 individuals between ages 40-to 69-years old enrolled between 2006 and 2010 (24, 25). The baseline visit, defined as the date of assessment center visit, included questionnaires and interviews that captured sociodemographic and health status, physical measurements, and phlebotomy measurements. Detailed information and measurements for each individual were recorded.

Methods for follow-up included linking individuals to national hospital inpatient and outpatient records, death registries, and primary care diagnoses (25). We excluded individuals with a history of CHD at baseline. Detailed exclusion criteria are summarized in **Supplementary Table 1**. In total, 387,832 individuals were considered. Age was calculated from the date of assessment center visit (data field 53) and the date of birth (data field 33). We adjusted our analysis for sex (data field 31). Measured risk factors considered in our analysis included body mass index (BMI, data field 21001), systolic blood pressure (SBP, data field 4080), smoking status (data field 20116), high-density lipoprotein cholesterol (HDL cholesterol, data field 30760), and total cholesterol (data field 30690). Analyses included all UK Biobank participants with relevant information and did not exclude individuals of non-European ancestry. All UK Biobank participants were included because the goal of the study was not to assess the prediction performance of PS per se, which was expected to be lower in individuals of non-European ancestry than in individuals of European ancestry. Rather, the goal of the study was to examine the association between PS and ACS across different clinical conditions. In a sensitivity analysis, we examined this association separately for each ancestry group defined using the same approach as used in the Pan-UK Biobank project (https://pan.ukbb.broadinstitute.org/).

FinnGen is a prospective cohort study that includes approximately 224,000 individuals representing about 3% of the Finnish adult population (21). Individuals were linked to national hospital discharge, death, and medication reimbursement registries. The baseline visit was defined as the date of DNA sample collection. Similar exclusion criteria were applied to FinnGen, in which 177,876 individuals were considered.

### Clinical conditions and outcomes

In the UK Biobank, clinical conditions were defined by the International Classification of Diseases, Tenth Revision (ICD-10) obtained from the Hospital Episode Statistics (HES) database which covers inpatient (1997–present) and outpatient (2003–present) admissions to hospitals. In individuals who developed ACS, we only considered clinical conditions that were diagnosed before baseline and at least 30 days prior to an ACS event to remove clinical conditions that may be captured or confounded by an ACS event. We excluded clinical conditions diagnosed after baseline to reflect a clinical setting in which prediction of ACS is assessed using past medical history. We excluded clinical conditions involving external causes of morbidity and mortality and factors influencing health status and contact with health services (ICD-10 chapters XX and XXI). In the case where an individual received multiple diagnoses for the same clinical condition, only the date of the first diagnosis was considered. We only considered clinical conditions with at least 10 co-occurrences with ACS to improve statistical power. Detailed exclusion criteria are summarized in **Supplementary Table 1**. ACS was algorithmically defined with ICD-10 codes capturing unstable angina (I20.0), acute myocardial infarction (I21), and subsequent ST elevation (STEMI) and non-ST elevation (NSTEMI) myocardial infarction (I22).

In FinnGen, clinical conditions were based on inpatient (1995-present) and outpatient (1998-present) admissions to hospitals. The same definition of ACS and exclusion criteria from the UK Biobank were used in FinnGen. From the UK Biobank, 9,080 cases of ACS and 405 ICD-10 codes were considered after exclusion. In FinnGen, 6,430 cases of ACS and 645 ICD-10 codes were considered after exclusion. Baseline characteristics of each cohort are summarized in **Table 1**.

**Table 1:** Summary of baseline characteristics of cohorts from UK Biobank and FinnGen.

|  | UK Biobank | FinnGen |
| --- | --- | --- |
| Number of individuals without ACS at baseline | 387832 | 177876 |
| Number of ACS cases (%) | 9080 (2.34) | 6430 (3.61) |
| Number angina cases (%) | 5477 (1.41) | 8326 (4.68) |
| Average follow-up time in years (SD) | 11.97 (1.36) | 7.18 (8.29) |
| Average age (SD) | 56.47 (8.09) | 51.29 (17.33) |
| Number of females (%) | 210340 (54.23) | 100929 (56.74) |
| Number of statin users (%) | 65419 (16.87) | 53886 (30.29) |
| Number of smokers (%) | 174763 (45.06) | - |
| Average body mass index (SD) | 27.38 (4.75) | - |
| Average systolic blood pressure (SD) | 139.8 (19.64) | - |
| Average high-density lipoprotein cholesterol (SD) | 1.45 (0.38) | - |
| Average total cholesterol (SD) | 5.71 (1.13) | - |

### Polygenic scores

PS was constructed using PRS-CS. Briefly, PRS-CS is a polygenic prediction method that uses Bayesian regression and infers posterior SNP effect sizes under continuous shrinkage priors using only GWAS summary statistics and an external linkage disequilibrium reference panel (26). PRS-CS was validated in several traits and common complex diseases, including CHD, and was robust and comparable to other well-established methods. To construct PS for individuals in the UK Biobank and FinnGen, we used summary statistics from the largest GWAS of CHD to date (20). This GWAS excluded individuals from the UK Biobank and FinnGen andincluded 60,801 cases and 123,504 controls. Case status was defined by an inclusive diagnosis of CHD. Summary statistics and the 1000 Genomes EUR reference panel were used to output weights using standard PRS-CS settings. Only HapMap3 variants were included. PS was calculated using PLINK 2.0 (27).

### Statistical analysis

First, we associated 405 clinical conditions diagnosed before baseline with ACS in the UK Biobank using Cox regression survival models, modeling clinical conditions as time-varying covariates. Baseline was defined as the date of assessment center visit. The end of follow-up date was defined as an ACS event, death, or our pre-defined end of follow-up date, March 31, 2020. Models were adjusted for age at baseline, sex, and principal components to control for ancestry. We adjusted subsequent models for measured risk factors, including BMI, SBP, smoking status, HDL cholesterol, and total cholesterol. This analysis was replicated in FinnGen with 645 clinical conditions. All models were evaluated for statistical significance after multiple testing correction.

Next, we looked at the interaction between clinical conditions and PS in models testing for the association with ACS. Only clinical conditions that were associated with ACS after multiple testing correction were considered (UK Biobank p-value<1.23×10^−4^, FinnGen p-value<7.75×10^−5^). Among clinical conditions with significant interactions, we explored whether there were differences in the prediction performance between individuals with and without these clinical conditions. We looked at the effect sizes (hazard ratio), discrimination (Harrell’s C-index), and net reclassification index (NRI) to evaluate the association and the prediction performance (28, 29). To calculate NRI, we used four risk categories of 0-5%, 5-7.5%, 7.5-20%, and >20% based on the 10-year risk for ACS including measured risk factors (12, 30, 31).

## Results

### Several clinical conditions are associated with acute coronary syndrome

In the UK Biobank, we explored the association between 405 clinical conditions diagnosed before baseline and 9,080 cases of ACS and identified 80 clinical conditions that were significantly (p-value<1.23×10^−4^) associated with ACS after multiple testing correction (**Figure 1**). The 80 associated clinical conditions had an average hazard ratio of 2.4 for ACS. Some of these associations were conventional (e.g., stable angina pectoris (SAP), type 2 diabetes mellitus) and others were unconventional (e.g., diaphragmatic hernia, inguinal hernia). Adjustment for measured risk factors modestly reduced the association, but statistical significance was retained for all 80 clinical conditions. **Supplementary Table 2** includes all 80 clinical conditions that were associated with ACS before and after adjustment for measured risk factors. This analysis was replicated in FinnGen, in which we explored the association between 645 clinical conditions and 6,430 cases of ACS and identified 71 clinical conditions that were significantly (p-value<7.75×10^−5^) associated with ACS after multiple testing correction (**Supplementary Figure 1**). The 71 associated clinical conditions had an average hazard ratio of 2.1 for ACS. **Supplementary Table 3** includes all 71 clinical conditions that were associated with ACS. Thirty-three clinical conditions were associated with ACS in both the UK Biobank and FinnGen. For these clinical conditions, we observed good consistency (R^2^=0.65) between the association observed in the UK Biobank and FinnGen, suggesting the generalizability of these associations across two large-scale biobanks (**Supplementary Figure 3**).

**Figure 1:**
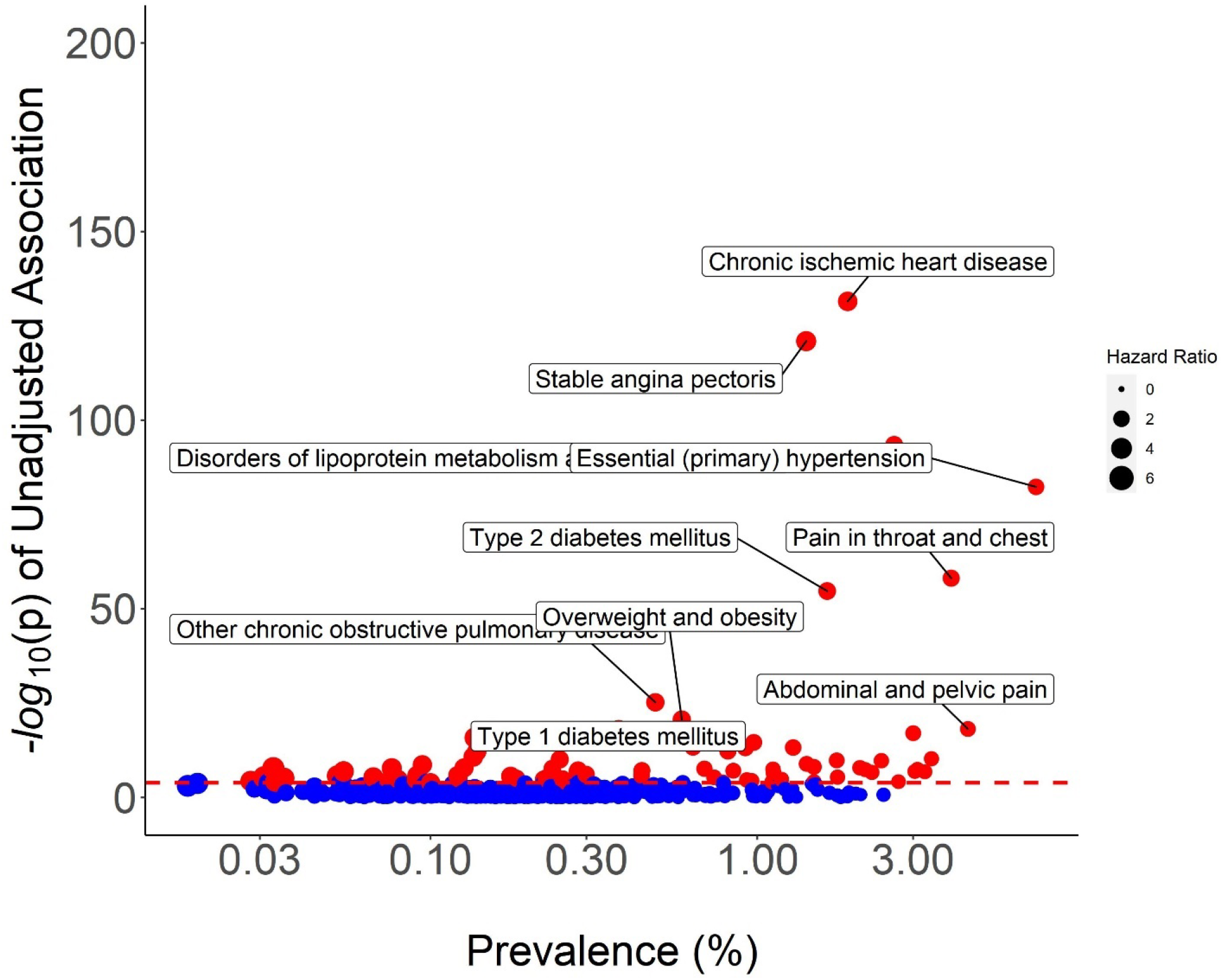
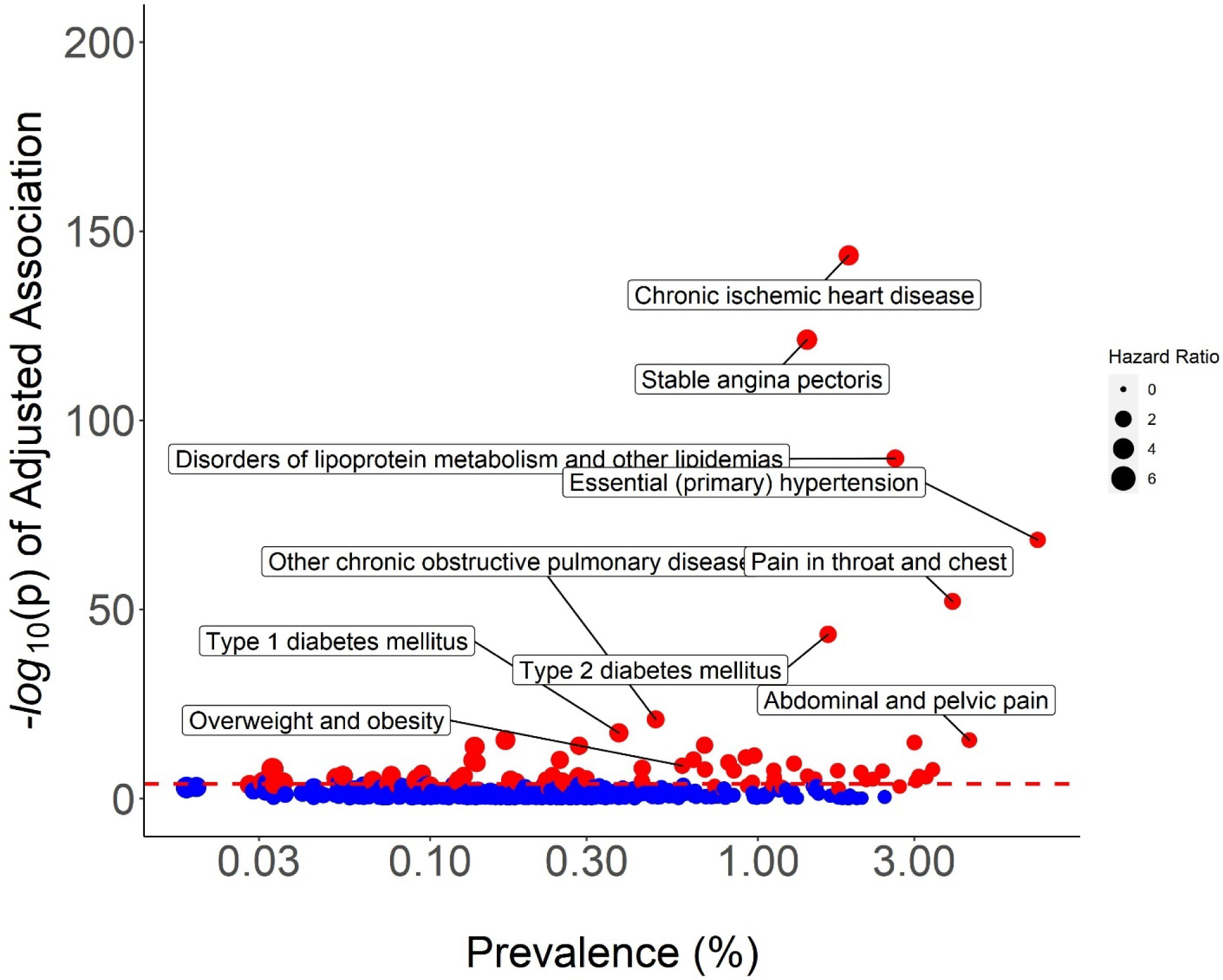
Association between 405 clinical conditions and 9,080 cases of ACS unadjusted (top) and adjusted (bottom) for measured risk factors from the UK Biobank. We used time-varying covariate Cox survival models adjusted for age, sex, and principal components. Measured risk factors included BMI, SBP, smoking status, HDL cholesterol, and total cholesterol. Red dotted line is the significance threshold after multiple testing correction (p-value<1.23×10^−4^). Top 10 clinical conditions are labeled.

### Most clinical conditions do not impact association between polygenic scores and acute coronary syndrome except for stable angina pectoris

First, we confirmed that PS was associated with ACS in the UK Biobank (HR for 1 SD increase in PS=1.49 [95% CI: 1.46–1.53], p-value=1.36×10^−294^) and FinnGen (HR for 1 SD increase in PS=1.44 [95% CI: 1.40–1.47], p-value=1.71×10^−166^). Next, we aimed to understand the value of measuring PS in individuals with or without clinical conditions associated with ACS and how the association between PS and ACS changed given the diagnosis of certain clinical conditions. We tested for the interaction between PS and 80 clinical conditions that were associated with ACS in the UK Biobank and identified a significant interaction (p-value=2.87×10^−8^) between SAP and PS (**Figure 2**). Here, SAP was defined as angina pectoris with documented spasm, other forms of angina pectoris, and unspecified angina pectoris (ICD-10 codes I20.1, I20.8, I20.9). This was the only interaction (p-value=1.38×10^−6^) that was replicated in FinnGen (**Supplementary Figure 2**). In the UK Biobank, there were 5,477 cases of SAP (prevalence=1.41%), and in FinnGen, there were 8,326 cases of SAP (prevalence=4.68%). There was also a significant interaction between PS and type 2 diabetes mellitus (p-value=6.01×10^−6^) in the UK Biobank, but this interaction was not significant after multiple testing correction in FinnGen (p-value=5.00×10^−2^).

**Figure 2:**
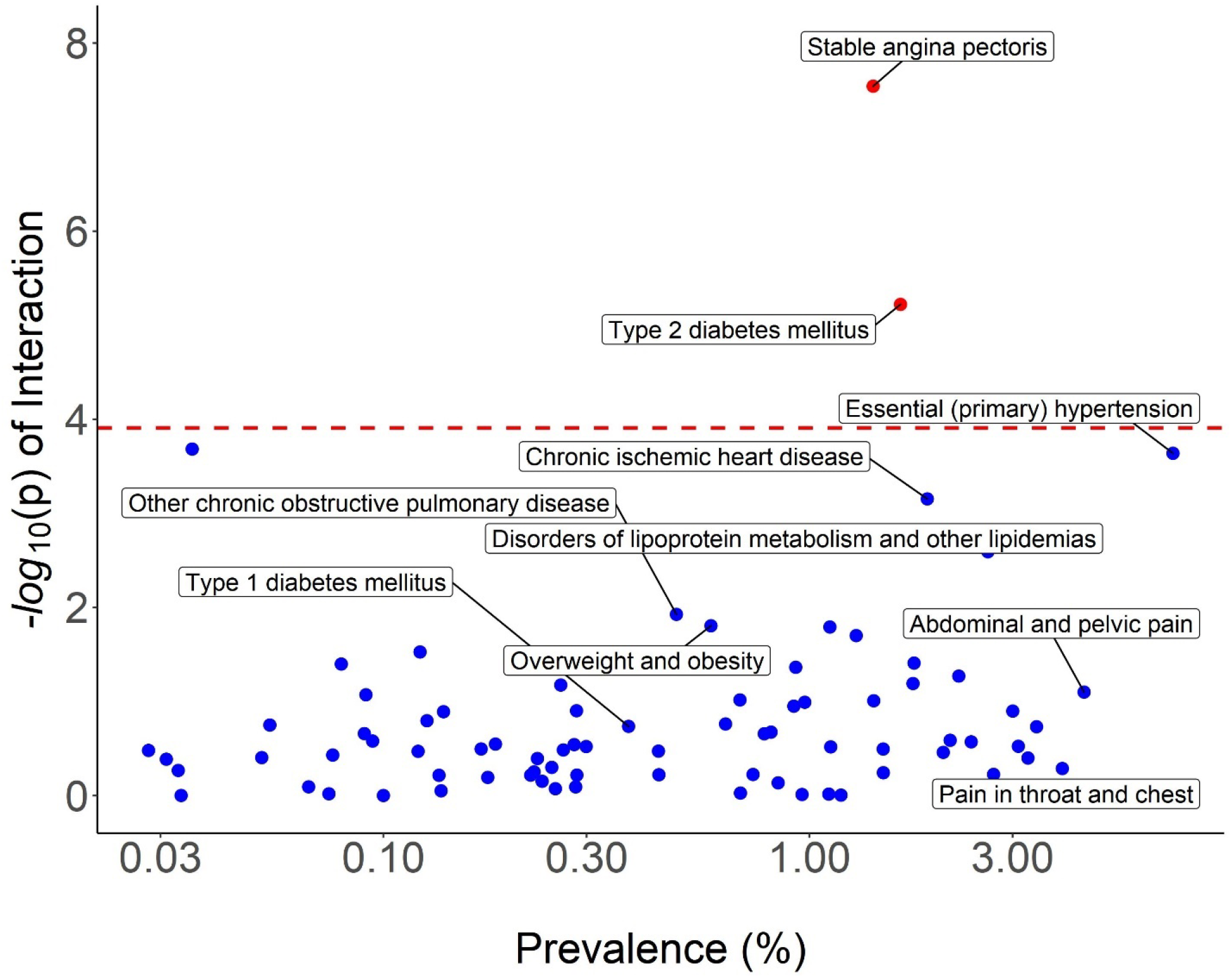
Interaction between PS and 80 clinical conditions significantly (p-value<1.23×10^−4^) associated with ACS after multiple testing correction from the UK Biobank. Red dotted line is the significance threshold (p-value<1.23×10^−4^). Top 10 clinical conditions are labeled. There was a significant interaction between SAP and PS in association with ACS (p-value=2.87×10^−8^) that was replicated in FinnGen (p-value=1.38×10^−6^). There was also a significant interaction between SAP and type 2 diabetes mellitus (p-value=6.01×10^−6^) in the UK Biobank, but this interaction was not significant after multiple testing correction in FinnGen (p-value=5.00×10^−2^).

In the UK Biobank, individuals with SAP had a significantly reduced risk for ACS (HR for 1 SD=1.163 [95% CI: 1.082–1.251]) compared to individuals without SAP (HR for 1 SD=1.531 [95% CI: 1.497–1.565]) (**Figure 3**). In a model including measured risk factors in the UK Biobank, results remained consistent (SAP HR=1.159 [95% CI: 1.078–1.246], no SAP HR=1.488 [95% CI: 1.455–1.522], interaction p-value=2.05×10^−7^). This observation was replicated in FinnGen, and individuals with SAP had a significantly reduced risk for ACS (HR for 1 SD=1.247 [95% CI: 1.173–1.326]) compared to individuals without SAP (HR for 1 SD=1.448 [95% CI: 1.407–1.490]).

**Figure 3:**
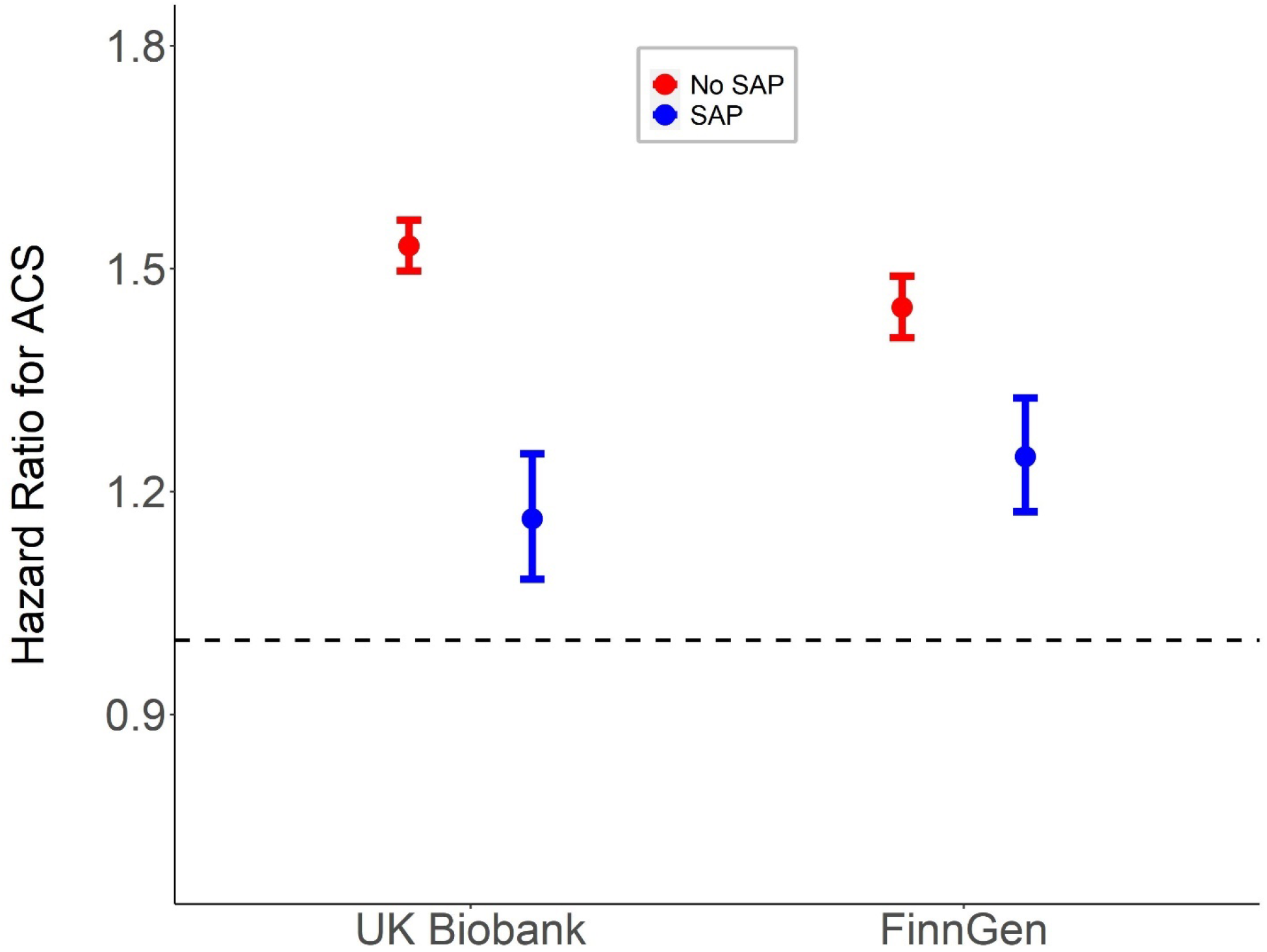
Hazard ratios for association between PS and ASC in individuals with and without SAP. In the UK Biobank, individuals with SAP had a hazard ratio of 1.163 [95% CI: 1.082– 1.251] and individuals without SAP had a hazard ratio of 1.531 [95% CI: 1.497–1.565]. In FinnGen, individuals with SAP had a hazard ratio of 1.247 [95% CI: 1.173–1.326] and individuals without SAP had a hazard ratio of 1.448 [95% CI: 1.407–1.490].

We also noticed that the association between measured risk factors and ACS were generally weaker among individuals with SAP than individuals without SAP (**Supplementary Table 4**). For example, there was no association between total cholesterol and ACS in individuals with SAP (HR=0.96 [95% CI: 0.89–1.03], p-value=0.247), but there was a significant association in individuals without SAP (HR=1.11 [95% CI: 1.09–1.13], p-value=6.43×10^−28^)

### Sensitivity analyses

We performed several sensitivity analyses. First, because PS has been shown to have age-dependent effects (21) and individuals with SAP tended to be older and include more males than individuals without SAP, we performed an age- and sex-matched analysis to test whether differences in age or sex explained the differential association of PS and ACS. After matching age and sex distributions in individuals with and without angina, the interaction between SAP and PS remained significant in both the UK Biobank (p-value=1.51×10^−3^) and FinnGen (p-value=1.16×10^−2^).

Second, because many well-established guidelines suggest statin interventions for individuals with SAP, we adjusted our analysis for statin use to explore whether statin use in individuals with SAP lowered the association with ACS. Statin use in the UK Biobank was self-reported and statin use in FinnGen was defined using high-quality prescription registries. In the UK Biobank, 82.1% of individuals with SAP used statins and 15.9% of individuals without SAP used statins. In FinnGen, 81.7% of individuals with SAP used statins and 27.8% of individuals without SAP used statins. The interaction between SAP and PS remained significant after adjusting for statin use in both the UK Biobank (p-value=1.20×10^−10^) and FinnGen (p-value=3.77×10^−4^). In the UK Biobank, the interaction between SAP and PS remained significant (p-value=7.39×10^−4^) after removing statin users.

Third, we increased the time window between a diagnosis of SAP and an ACS event to reduce the chances of capturing the same underlying cardiovascular event. Increasing the time window from 30 days to 300 and 500 days only modestly reduced the interaction between SAP and PS in the UK Biobank (300 day p-value=5.14×10^−11^, 500 day p-value=3.76×10^−11^) and FinnGen (300 day p-value=5.83×10^−4^, 500 day p-value=1.86×10^−3^). Average and median time between an SAP diagnosis and an ACS event was 9.1 and 8.9 years, respectively, in the UK Biobank. Similarly, average and median time was 6.7 and 5.7 years, respectively, in FinnGen. This suggested that the cases of SAP captured in our study were diagnosed earlier and independent of ACS.

Fourth, we examined the interaction between SAP and ACS in different ancestry groups. Our cohort from the UK Biobank included individuals of African (AFR, n = 5,543), Ad Mixed American (AMR, n = 820), Central and South Asian (CSA, n = 6,976), East Asian (EAS, n = 2,196), European (EUR, n = 339,771), and Middle Eastern (MID, n = 1,258) ancestry. In CSA (ACS cases = 306, SAP diagnoses = 194, p-value = 1.14×10-2) and EUR (ACS cases = 7,969, SAP diagnoses = 4,810, p-value = 2.36×10-11), the interaction between SAP and PS remained significant. Other ancestry groups, which included less than 100 diagnoses of SAP and 100 cases of ACS, were not analyzed due to the lack of statistical power.

### Prediction performance of polygenic scores differs between individuals with and without stable angina pectoris

We examined Harrell’s C-index to evaluate the prediction performance of PS (**Figure 4**). In the UK Biobank, individuals with SAP had a C-index for PS of 0.606 [95% CI: 0.595–0.616] and individuals without SAP had a C-index for PS of 0.747 [95% CI: 0.743–0.750] when considering models adjusted for age, sex, principal components, BMI, SBP, smoking status, HDL cholesterol, and total cholesterol. These findings were replicated in FinnGen, and individuals with SAP had a C-index of 0.665 [95% CI: 0.655–0.674] and individuals without SAP had a C-index of 0.825 [95% CI: 0.821–0.830]. The lower C-index in individuals with SAP was explained by the lower association between established cardiovascular risk factors, including age, and ACS in these individuals.

**Figure 4:**
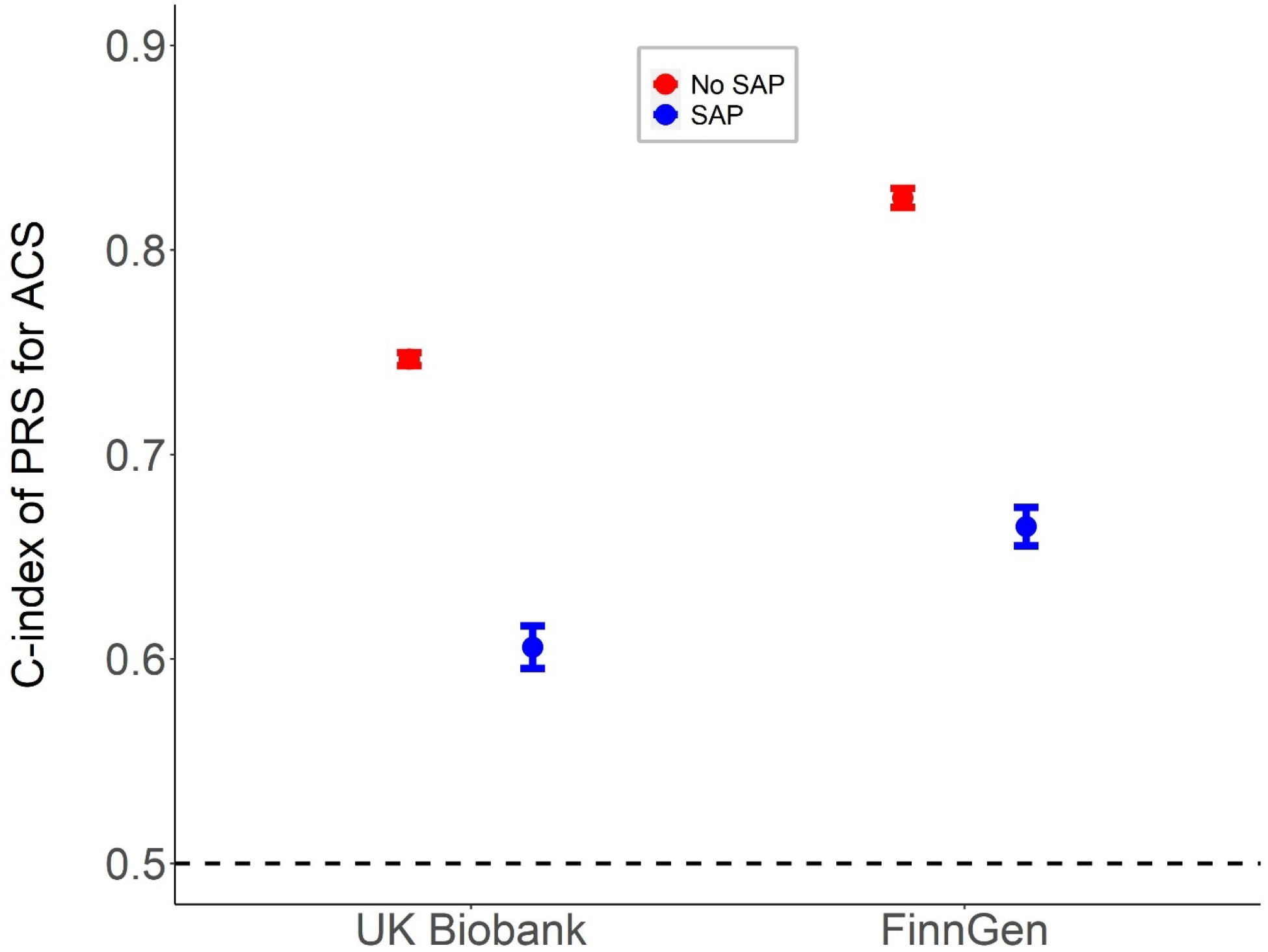
Difference in discrimination for ACS, measured by Harrell’s C-index. In the UK Biobank, individuals with SAP had a C-index of 0.606 [95% CI: 0.595–0.616] and individuals without SAP had a C-index of 0.747 [95% CI: 0.743–0.750]. In FinnGen, individuals with SAP had a C-index of 0.665 [95% CI: 0.655–0.674] and individuals without SAP had a C-index of 0.825 [95% CI: 0.821–0.830]. The lower C-index in individuals with SAP is explained by the lower association between established cardiovascular risk factors, including age, and ACS in these individuals.

Because it is easier to obtain a higher improvement in predictive performance (ΔC-index) when the baseline model has a lower prediction performance (32, 33), we expected the improvement in C-index due to PS to be higher in individuals with SAP. Nevertheless, the addition of PS to measured risk factors increased the C-index more in individuals without SAP (ΔC-index=0.021) than in individuals with SAP (ΔC-index=0.010). In FinnGen, PS increased the C-index by 0.012 in individuals without SAP and by 0.013 in individuals with SAP. Lastly, we calculated the NRI to evaluate the improvement in reclassification when adding PS to the baseline prediction performance of measured risk factors (**Supplementary Table 5**). Addition of PS significantly improved the prediction performance in individuals without SAP (categorical NRI=0.116, p-value<1.00×10^−3^; continuous NRI=0.315, p-value<1.00×10^−3^) but not in individuals with SAP (NRI=0.038, p-value=0.187; continuous NRI=0.113, p-value=3.41×10^−3^). Overall, the NRI was higher among individuals without SAP compared to individuals with SAP.

## Discussion

In this study, we performed an unbiased, comprehensive evaluation of the association between diverse correlated clinical conditions and ACS across two large-scale biobanks. Moreover, by exploring the impact of clinical conditions on the association between PS and ACS, we provided context for the use of PS in a clinical setting for prediction of ACS.

First, we found that a large number of clinical conditions were associated with ACS, independent of measured risk factors. Using clinical conditions to predict risk for ACS may be more convenient than using measured risk factors because this information is easily accessed and obtained from electronic health records and does not require in-person visits. In addition to well-established clinical risk factors, such as type 2 diabetes mellitus, that are routinely considered, additional clinical conditions that are associated with ACS may be taken into consideration when evaluating risk for ACS. Identification of such clinical conditions is critical to provide a comprehensive, well-informed assessment of risk for ACS.

Second, by using a hypothesis-free approach across 80 clinical conditions associated with ACS, we found that the association between PS and ACS was consistent across individuals with or without most of these clinical conditions. Diagnosis of a clinical condition did not change the association between PS and ACS, suggesting that clinical conditions do not impact the utility of PS. Previous work has shown that different groups of individuals may benefit more from measuring PS than other groups. For example, previous work has shown that PS was more predictive of ACS among individuals who had never smoked (34). However, in our study, we generally found that the diagnosis of a clinical condition did not affect the association between PS and ACS. The only differential association that was found in both the UK Biobank and FinnGen was a diagnosis of SAP, where SAP was significantly more associated with ACS in individuals without a previous diagnosis of SAP than in individuals with a previous diagnosis of SAP.

There are at least two possible explanations for this phenomenon. First, progression of ACS occurs with or without the presentation of SAP. It is possible that individuals who develop ACS with or without prior SAP are clinically and genetically distinct. It is widely accepted that ACS varies in clinical presentation, and other studies have explored the significance of a diagnosis of SAP before ACS (35-38). For example, other studies have found that individuals with prior SAP had more extensive atherosclerosis and a greater prevalence of clinical risk factors, such as hypertension, than individuals without prior SAP who developed ACS (39, 40). In another study, myocardial infarction was the most common first clinical presentation in individuals without prior SAP who developed ACS (35). Other studies have also found that individuals with prior SAP had a more favorable prognosis compared to individuals without prior SAP who developed ACS (39, 41). In our study, we found a differential association between PS and ACS in individuals with and without SAP, further suggesting that these individuals may be clinically and genetically distinct. Taken together, there may be different pathways of progression of ACS that may warrant different assessments and interventions.

Second, a diagnosis of SAP may be synonymous to a diagnosis of CHD. In a clinical setting, individuals with SAP often have already started developing CHD. Subsequently, individuals with SAP may already have a higher risk for ACS, and PS may not be informative. As a result, PS may lose its prediction performance in individuals who have already been diagnosed with SAP. This observation is supported by other studies that have shown that prediction of secondary CHD events by PS is attenuated compared to that of primary CHD events (42).

An open matter that may explain this phenomenon is index bias. Index bias may occur in studies that select patients based on the occurrence of an index event, and as a result, measured risk factors may have less of an effect on recurrent events (43-45). In our study, SAP may be an index event that decreased the prediction performance of PS in individuals with SAP compared to individuals without SAP. In other words, because ACS is dependent on multiple factors, such as underlying chronic conditions and measured risk factors, conditioning on a diagnosis of SAP creates a dependence between such factors. As a result, individuals with SAP do not require the same burden of increased measured risk factors and PS to develop ACS. As mentioned previously, individuals with SAP are more likely to have underlying chronic conditions, such as extensive atherosclerosis. This may entail that measured risk factors and PS do not need to have a large association with ACS to confer the same risk, due to the presence of such underlying chronic conditions. Indeed, our study showed that measured risk factors and PS were less associated with ACS in individuals with SAP, reflecting other studies that have also shown that individuals with SAP often have less associated measured risk factors (35). Thus, PS becomes less predictive of ACS in individuals with SAP than in individuals without SAP.

A few limitations of this study must be considered. First, definition of ACS and clinical conditions were based on diagnostic codes derived from national health registries.

Underreporting and incorrect use of diagnostic codes in a clinical setting are inherent to these types of data. Nevertheless, we obtained similar results in two large-scale biobanks from the United Kingdom and Finland. Additional evidence from other studies also support the quality of this type of data (46-48). Future studies are needed to fully assess the validity of using registry-based clinical conditions across large-scale biobanks from different countries, but our study supports the feasibility of cross-nation registry-based analyses. Second, our PS represents a “snapshot” of current research in genetic scores. PS is inherently tied to the study used to construct the genetic scores, and as larger GWAS are conducted, these genetic scores will necessarily improve and become more predictive. Therefore, our results represent a lower bound of prediction performance of PS, and it is likely that additional studies that improve genetic scores will improve the prediction performance of PS.

## Conclusion

Using an unbiased, comprehensive approach, we identified clinical conditions that were associated with ACS, independent of measured risk factors. Overall, we found that the association between PS and ACS remained consistent across many clinical conditions, suggesting that the utility of PS for prediction of ACS is largely independent of previous clinical conditions experienced by the patient. Nevertheless, we found one exception: individuals without SAP had a greater association between PS and ACS than individuals with SAP. In conclusion, we contextualized the use of PS in a clinical setting and showed that a PS for ACS may be more appropriate among asymptomatic individual than symptomatic individuals with clinical suspicion for CHD.

## Data Availability

UK Biobank data are available by request at https://www.ukbiobank.ac.uk/using-the-resource/. FinnGen data are available by request at https://www.finngen.fi/en/access_results.

## Acknowledgements

UK Biobank analyses were conducted under application 31063. We acknowledge all study participants for their generous participation in the UK Biobank and FinnGen. This work was supported by the Stanford University Undergraduate Research Student Grant.

## Notes

### Competing Interest Statement

The authors have declared no competing interest.

### Author Declarations

The UK Biobank was approved by the National Health Service National Research Ethics Service and all participants provided written informed consent. FinnGen was approved by the Coordinating Ethics Committee of the Helsinki and Uusimaa Hospital District.

